# Genome Alert!: a standardized procedure for genomic variant reinterpretation and automated genotype-phenotype reassessment in clinical routine

**DOI:** 10.1101/2021.07.13.21260422

**Authors:** Kevin Yauy, François Lecoquierre, Stéphanie Baert-Desurmont, Detlef Trost, Aicha Boughalem, Armelle Luscan, Jean-Marc Costa, Vanna Geromel, Laure Raymond, Pascale Richard, Sophie Coutant, Mélanie Broutin, Raphael Lanos, Quentin Fort, Stenzel Cackowski, Quentin Testard, Abdoulaye Diallo, Nicolas Soirat, Jean-Marc Holder, Nicolas Duforet-Frebourg, Anne-Laure Bouge, Sacha Beaumeunier, Denis Bertrand, Jerome Audoux, David Genevieve, Laurent Mesnard, Gael Nicolas, Julien Thevenon, Nicolas Philippe

## Abstract

Numerous countries have set up population genomics plans, allowing an unprecedented growth in the ability of interpreting variants in human diseases. Retrospective interpretation of sequenced data in the light of the current literature is a major concern of the field. Moreover, such reinterpretation is manual and both the human resources and the variable operating procedures are main bottlenecks.

This work describes the Genome Alert! standardized procedure. This open-source method automatically reports changes with potential clinical significance in variant classification between releases of the ClinVar database. Using ClinVar submissions across time, this method assigns gene-disease associations validity category. Genome Alert! was assessed on a retrospective 29 months multicentric series of 5,959 consecutive individuals screened by targeted or exome sequencing.

Between July 2017 and December 2019, the retrospective analysis of ClinVar submissions revealed a monthly median of 1,247 changes in variant classification with potential clinical significance and 23 new gene-disease associations. Reexamination of 4,929 targeted sequencing files highlighted 45 changes, which 89% classifications were expert validated, leading to four additional diagnoses. Genome Alert! gene-disease association catalog provided 75 high-confidence associations not available in the OMIM morbid list, where 20% became OMIM morbid 8 months later. Over 356 negative exome sequencing data that were reannotated for variants in these 75 genes, this elective approach led to a new diagnosis.

Genome Alert! (https://genomealert.univ-grenoble-alpes.fr/) enables the systematic and reproducible reinterpretation of acquired sequencing data in a clinical routine with a limited human resource impact.

**Graphical abstract:** 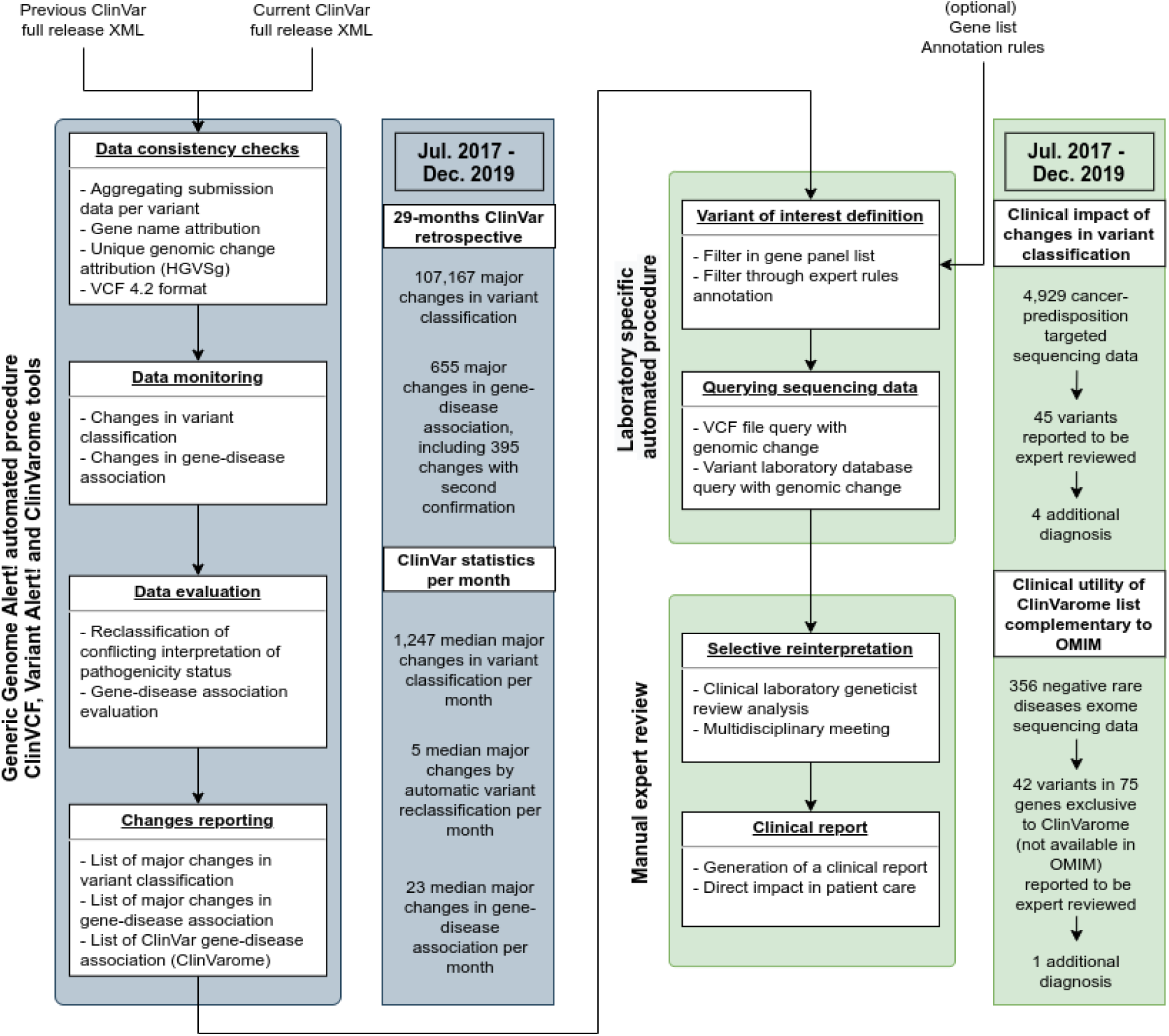

## Introduction

Genetic tests are increasingly prescribed and included in the healthcare pathways for diverse clinical indications ^1,2^. Several countries have developed population genomics organizations that are revolutionizing medical practices ^3,4^. However, many of these genomic analyses remain inconclusive, due to limitations in genomic and medical knowledge available at the time of analysis.

The ACMG-AMP recommendations for variant classification aim at standardizing variant interpretation practices in genomic centers, in the context of medical interpretation ^5^. Recently, tools have been published to automatically classify genomic variants, based on these recommendations ^6,7,8^. Meanwhile, the evolving medical knowledge and the rapid adoption of clinical genome sequencing has influenced the standard practices and created additional needs. A current and major preoccupation in this field is the definition of standards for periodic and prospective reanalysis of existing sequencing data. Indeed, reanalyzing existing genomic data improves diagnostic yield (7% increase per year) ^9,10^.

In practice, such an in-depth reinterpretation is mainly manual, time-consuming, with major bottlenecks such as human and funding resources or lack of consistency between centers. Clinical recommendations from the American and European Societies of Human Genetics reinforce the need for a standardized and automated approach to the reinterpretation of genomic analyses ^11,12,13,14^. Some companies offer paid “black box” services, with poorly detailed methods that can’t be reproduced ^15,16^. Clinical knowledge of rare diseases is contained in expert-curated databases (such as Online Mendelian Inheritance in Men or OMIM ^17^ or ClinGen ^18^), peer-reviewed medical literature, and information sharing between health practitioners through community-based platforms (as MatchMaker Exchange ^19^ or ClinVar ^20^). Reliability and exhaustiveness of information vary widely across these data sources. Furthermore, the careful monitoring of clinical knowledge by every laboratory represents an organizational challenge for a prospective re-analysis of acquired data. To enable a systematic, reproducible, and prospective genome interpretation, a collaborative approach to clinical knowledge aggregation combined with automated medical knowledge monitoring and curation is needed.

The main community-based repository of genomic knowledge is ClinVar (https://www.ncbi.nlm.nih.gov/clinvar/), a shared variant interpretation database that featured 1 million submissions in 2020. ClinVar is updated weekly with several thousands of modifications of variant classifications that could impact the diagnostic yield of previous analyses. There is currently no monitoring system that can highlight these changes at the scale of the complete database. Besides variant classification, gene-phenotype association catalogs are crucial as they are commonly used to design phenotype-specific gene panels for dry-lab filtering and set the frontiers of the clinical genome analysis ^21 22^. Though not their primary purpose, variant-centered databases could also theoretically provide a complementary resource to gather gene-phenotype knowledge.

Here, we detail an automated method for the re-assessment of variant pathogenicity and genotype-phenotype associations through ClinVar follow-up. This procedure, called Genome Alert!, aims at performing a routine and systematic reinterpretation of existing genomic data. The procedure’s effectiveness was evaluated through a 29-months multicentric series (2018-2019) of 5,959 consecutive individuals screened by targeted sequencing (4,929 individuals with hereditary cancers) and exome sequencing (1,000 analysis including 356 undiagnosed individuals with suspected Mendelian disorders).

## Methods

### Genome Alert! standardized procedure

#### ClinVCF - a ClinVar quality processing method

Before comparing different versions of the same source, data consistency needs to be verified. This first step is based on ClinVCF tool and once every change has been tracked, data will be processed for the next step.

ClinVCF (***See web resources***) imports monthly updated ClinVar XML files (***see web resources***). The XML format was preferred over the VCF mainly because of better consistency and traceability across versions for the ClinVar Variation ID, the history of changes in each variant classification, and the additional genotype-phenotype data available in XML. For each variant entry, ClinVCF query the gene name in a two-step process. When a gene symbol is provided and semantically correct according to the NCBI RefSeq GFF, the gene symbol is retained. Otherwise, the variant entry is annotated by ClinVCF according to the NCBI GFF. Then, ClinVCF proposes a consensus classification according to ClinVar policies (aggregation of ACMG interpretations provided in submitted records per variant) and gathers additional information provided by submitters (e.g. clinical terms or disease name).

Lastly, ClinVCF considers an automatic reclassification of variants with at least four submissions and conflicting interpretations of pathogenicity status. Consensus classification according to ClinVar policies sets the conflicting interpretations of pathogenicity status when at least one conflict in submission is observed. Based on the provided classifications transformed from literal transcription (e.g Likely pathogenic) to class number (e.g. class 4), if >=4 submissions are available, a new consensus is proposed after outlier submissions removal according to the 1.5* Interquartile Range (IQR) Tukey method ^23^.

ClinVCF provides a three-tier reclassification confidence score detailed in **Figure S1**. We only reclassify variants from conflicting status to likely pathogenic or pathogenic status, with a default first tier confidence score.

To ascertain the robustness of this reclassification method, we have evaluated this automatic reclassification when adding a variant of unknown significance (VUS) in the data submissions. Adding noise can be considered as a defensive approach, in a similar idea to what is being used in deep learning ^24^. This test aims at verifying if the amount of data is sufficient to draw similar conclusions, in the event of an additional virtual VUS submission (second tier confidence). As some reclassifications only rely on likely pathogenic submissions, a definitive reclassification is performed only if at least one pathogenic submission is available (third tier confidence). As an output, ClinVCF writes a VCF v4.2 file adding the following annotations if an automatic reclassification is performed: proposed reclassification in CLNSIG, ClinVar conflicting interpretations of pathogenicity stats in OLD_CLNSIG, reclassification confidence score in CLNRECSTAT.

#### Variant Alert! - a variant knowledge monitoring tool

Variant Alert! tool aims at identifying changes in variant classification across two versions of the database. Changes were defined as (i) a modification in the classification of an existing variant, (ii) the creation or suppression of a variant entry.

A stratification of the consequences in a classification modification was proposed (**Table S1**). A major classification modification was defined as a change that may impact the clinical management of the patient (e.g. Unknown Significance to Likely Pathogenic status). A minor classification modification was defined as a change that may not impact the clinical management of the patient (e.g. Pathogenic to Likely Pathogenic status).

Variant Alert! writes two files : (i) the list of variants that were modified, added, or removed and (ii) the list of genes that were added or removed to the database. This gene list is notably used by ClinVarome.

#### ClinVarome - a method for automated gene-disease association evaluation

ClinVarome tool aims to periodically and automatically evaluate gene-disease association in the ClinVar database. To differentiate genes based on their clinical validity, the work from EMBL-EBI Gene2Phenotype ^25^, ClinGen ^18^, Genomic England PanelApp^26^ was first compared. Although theoretically comparable, their rationales and contents were partially overlapping and with conflicting classifications. To discriminate candidate genes from definitive gene-disease associations, we decided to use an unsupervised clustering model. Only the genes with at least one likely pathogenic or pathogenic variant (single nucleotide variant or indel affecting a single gene) in ClinVar were considered in a list called ClinVarome. As a consensus criterion, based on a ClinGen recommandation, we choose to assess the strength of a gene-disease association through the quantification of four variables: (i) count of likely pathogenic and pathogenic variants, (ii) highest variant classification (CLNSIG, likely pathogenic or pathogenic), (iii) highest ClinVar review variant confidence (CLNREVSTAT, from zero to four stars) and (iv) time interval between the first to the last pathogenic variant submission (replication of the gene-disease association event) ^27^. For these four variables, values were gathered through periodic monitoring of changes in the database following the ClinVCF and Variant Alert! tool procedures. Clustering variants according to these variables allowed us to define clusters of genes according to their clinical validity. The scikit-learn Agglomerative Clustering tool (parameters: euclidean affinity, ward linkage) was used and t-SNE representation (parameters: 2 components, perplexity 150, 2,000 iterations, and 1,000 iterations without progress) was performed.

### Genome Alert! comparison with public database resources

Monthly ClinVar full XML release data from 2017-06-20 to 2019-12-01 and from 2019-12-01 to 2020-08-03 were downloaded. From 2017-06-20 to 2019-12-01 data were used for the ClinVar retrospective and the sequencing analysis reinterpretation. The impact of changes were measured on gene groups based on the *in silico* gene panels and disease groups from the Genomics England PanelApp ^26^ API on 01-06-2020. Gene symbols gathered from multiple resources (OMIM, ClinVar, and PanelApp) were unified with their NCBI Gene ID (via NCBI RefSeq annotation). To evaluate the exhaustivity of ClinVar morbid gene knowledge, a comparison between the list of all ClinVar morbid genes named *ClinVarome* with the gold standard OMIM database morbid clinical gene list was performed ^17^. An OMIM gene is defined as a morbid clinical gene if at least one phenotype or disease syndrome was associated with the gene at that time.

ClinVar data from 2019-12-01 to 2020-08-03 were used to validate ClinVarome. Identification of the gene-phenotype morbid list was made through the OMIM morbid map list (downloaded on 2019-11-14 and 2020-08-24) via the OMIM API^17^.

### Study Design and Participants

To evaluate the clinical impact of **Genome Alert!**, we collected 5,929 consecutive sequencing data samples from 3 centers in France between July 2017 and December 2019 as part of their routine genetic investigation : (i) a variant database gathering all class 3 (unknown significance), class 4 (likely pathogenic) and class 5 (pathogenic) variants identified in a colon cancer-targeted sequencing (14 genes) sequenced in 2,540 individuals in the Rouen University Hospital, (ii) a cancer-targeted sequencing dataset of 2,389 individuals by the Cerba laboratory (66 genes) and (iii) exome sequencing data of individuals with developmental disorders, rare kidney diseases or other rare diseases, as following: 108 probands from the Rouen University Hospital, 477 probands (with 356 negative analysis) from the Cerba laboratory and 415 probands from the Eurofins Biomnis laboratory. Patient samples, together with a basic phenotype description and molecular diagnosis (when available), were anonymized. Patients or legal guardians provided informed written consent for genetic analyses in a medical setting.

Two main clinical evaluations were performed: (i) Variant-centered reanalysis, which aims at matching individuals that carry exact variants with potential clinical significance reported by Genome Alert! and (ii) Gene-centered reanalysis, which aims at matching individuals that carry predicted variants in high-confidence clinical genes referenced in ClinVarome and not in OMIM.

### Selection of variants with potential clinical significance

All sequencing data were systematically reinterpreted according to Genome Alert!’s report and compared to the initial variant interpretation. For targeted sequencing and exome reanalysis, genomic positions of variants with major changes in classification were queried in the existing patient’s variant calling files (Variant-centered analysis). For exome data, we performed a reanalysis of variants in VCF with the following criteria: (i) among 75 *ClinVarome* morbid genes not available in OMIM and with a second event of gene-disease validation (including a likely pathogenic or pathogenic variant with ClinVar review confidence >= 2 stars and a likely pathogenic or pathogenic variant entry subsequent to the initial entry), and (ii) variant not shared with another individual in the series, and (iii) sufficient sequencing quality (variant allele fraction > 25% and read depth > 20 reads), and (iv) rare in gnomAD^28^ population (frequency < 10^−5^ if heterozygous genotype or 10^−4^ if homozygous genotype), and (v) protein consequence among nonsense or frameshift or missense variants (missense are selected with CADD^29^ > 30 and MetaSVM^30^ = D) or splice variant (based on dbscsnv RF^31^ predicted impact score >0.6) (Gene-centered reanalysis).

## Results

### Monitoring ClinVar knowledge dynamics

To get insights into variant classification and gene-disease association as well as to estimate the amount of new clinically relevant information in the ClinVar database available through time, a retrospective analysis of ClinVar submissions over 29 months was performed (July 2017 (included) to December 2019). Of note, VCF genomic positions in ClinVar were introduced in July 2017 and probably are associated with the largest injection in the ClinVar database.

The number of variants with ACMG-AMP classification ^5^ increased from 144,943 to 491,838. Among modifications in the database, the count of major changes was 107,167 in ACMG-AMP classification and, among these, 103,615 resulted in a pathogenicity status previously unreported, while 3,552 resulted in the revocation of a previously established pathogenicity (Figure 1A). These changes varied significantly according to disease types between gene panels (according to Genomics England PanelApp ^26^), where top of them were oncogenetic panels. The panels presenting most of the changes per gene are presented in Figure 1B and Table S2. Clinical gene entries in ClinVar were also monitored. A median of 23 ClinVar morbid genes per month newly associated with Mendelian disease was observed (Figure 2).

**Figure 1.**
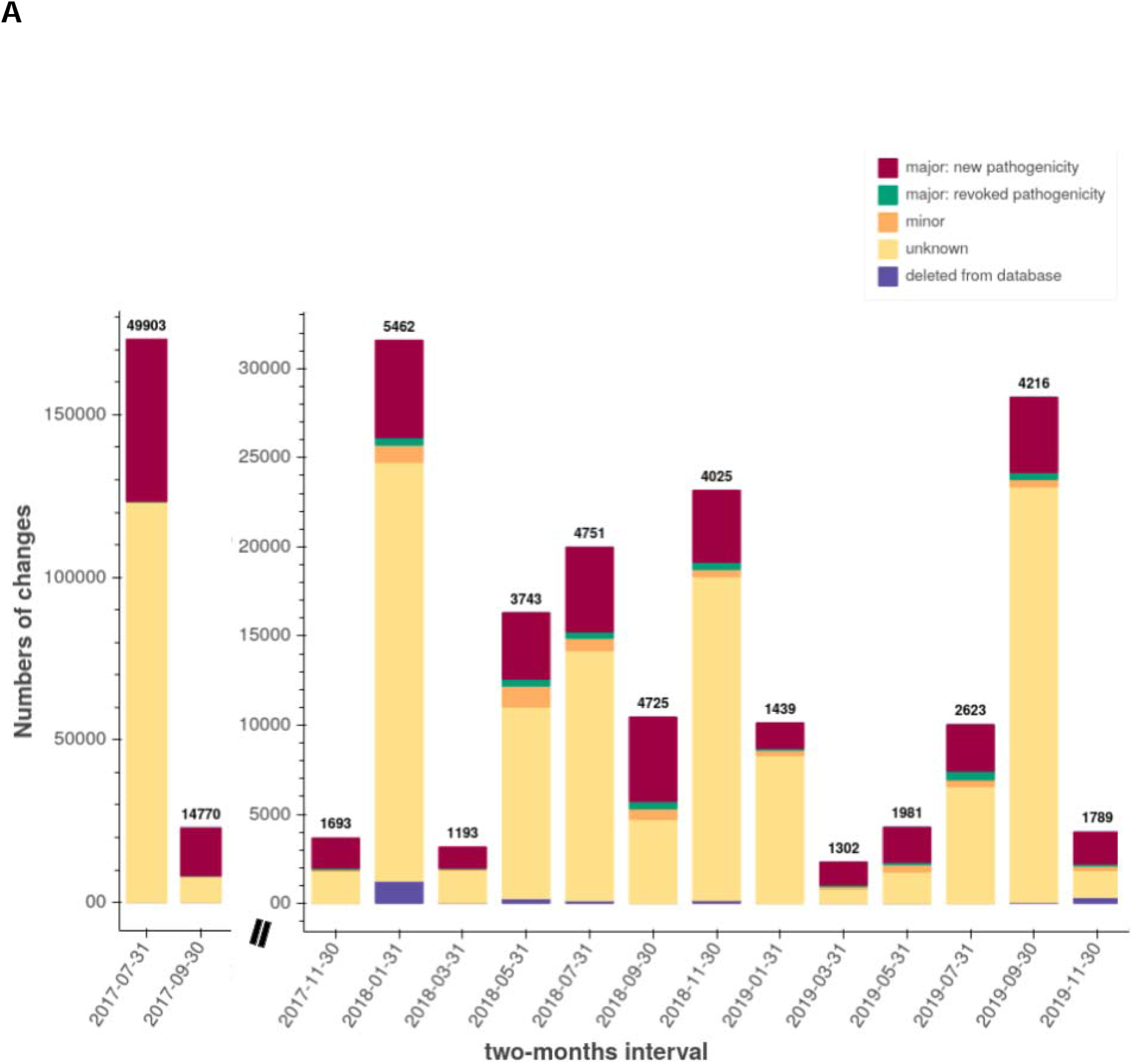
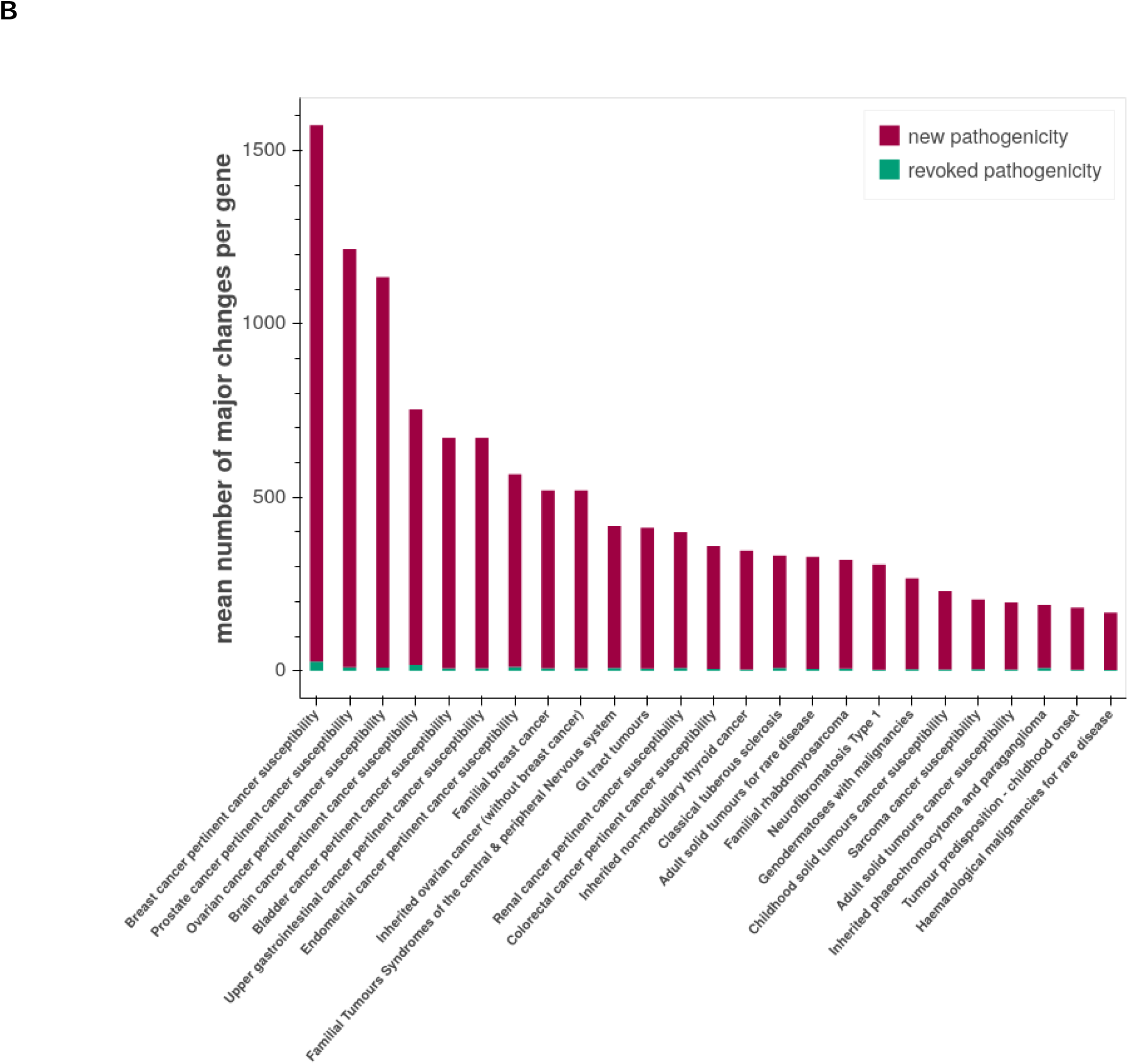
ClinVar variant classification monitoring between July 2017 to December 2019. A. Bar chart distribution every 2 months of changes in variant classification. The bar chart was split for a better readability. Bold numbers and dark red color identifies new (likely) pathogenic variant entries; Green = number of revoked (likely) pathogenic variants; Orange = number of minor change variants (e. g. pathogenic to likely pathogenic); Yellow = number of changes with no clinical impact; Purple = number of changes leading to variant disappearance. B. Bar chart of top panels with clinically significant changes per gene (major changes) Dark red color identifies (likely) pathogenic variant entries and Green for revoked (likely) pathogenic variants.

**Figure 2.**
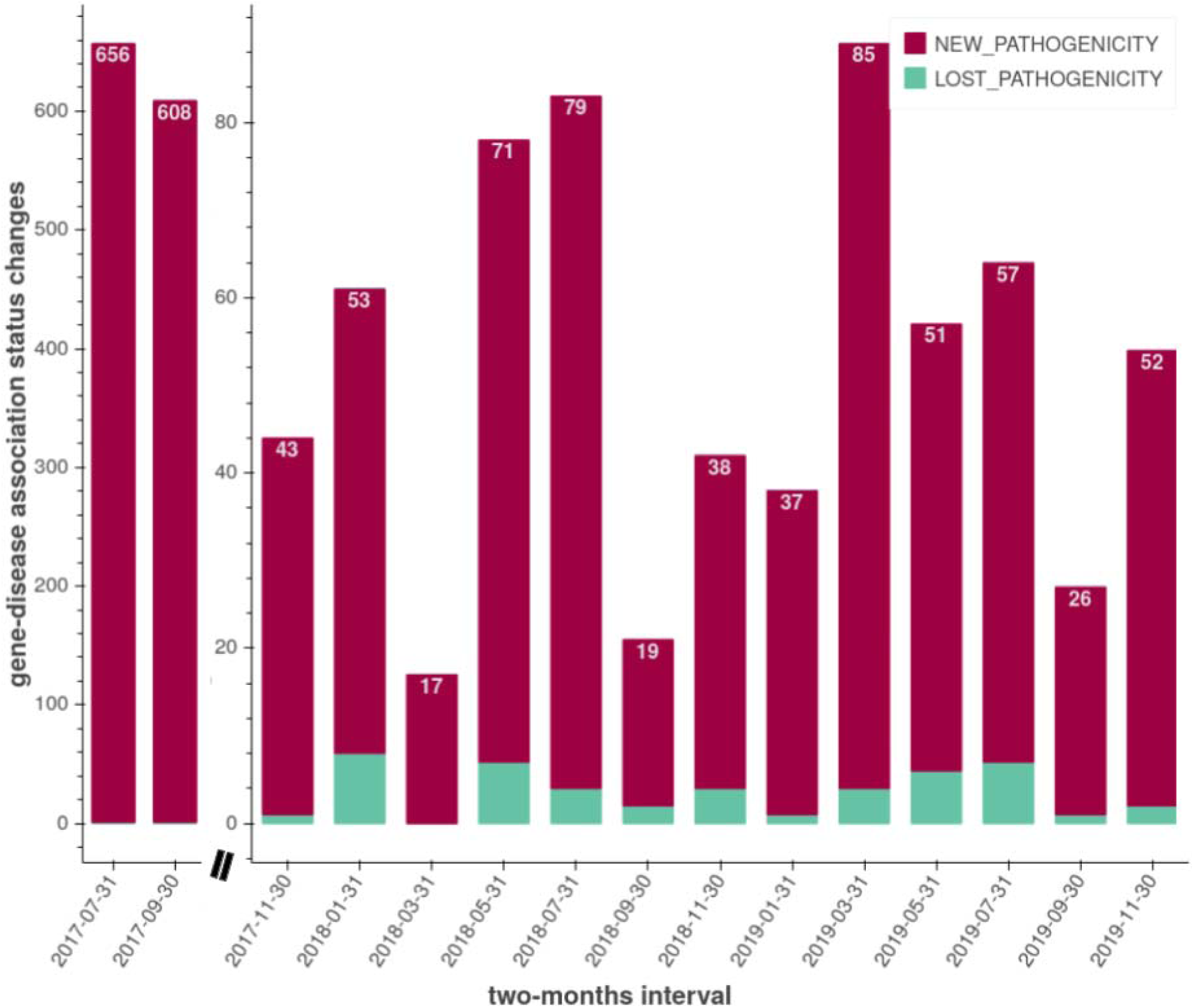
ClinVar clinical genes entries associated with new or deprecated mendelian disease (morbid status) distribution between December 2017 to December 2019. The bar chart was split for a better readability. Dark red = morbid genes entries (first variant with likely pathogenic or pathogenic status); Green = revoked morbid genes. White numbers = number of new morbid gene entries by two months.

### Evaluate ClinVar variant knowledge consistency

To evaluate the robustness of clinical variant information, the consistency of variant classification was explored (Table S3). Among 144,943 variants available in July 2017, 10,254 (7%) were reclassified between July 2017 and December 2019 meaning that we observed only a small portion of variants being reclassified over time. Among the 11,417 likely pathogenic variants, 1125 (9.94 %) variants were reclassified as benign, likely benign, of unknown significance, or conflicting interpretations of pathogenicity.

### Manual curation of automatic variant reclassification with conflicting interpretations

A common criticism of the ClinVar database is the misclassification of well known pathogenic variants. We observed that it was mostly due to a unique outlier submission. We evaluated our method to remove such outlier submissions. Among all the variants available in ClinVar in December 2019, 22,973 out of a total of 503,994 (4.5%) variants were classified with a conflicting interpretation of pathogenicity. Genome Alert! automatic reclassification method proposes to detect outlier submissions to suggest a consensus classification. This allowed the reclassification of 188 variants in 135 genes (Table S4 and Figure S1).

Variants automatically reclassified in cancer (n=9) and cardiogenetics disease (n=11) were presented to experts in the field. Of these 20 automatic reclassifications, 17 were confirmed as accurate by experts. Three variants remained of uncertain significance.

### Clinical impact of changes in variant classification

To assess the clinical impact of Genome Alert!’s changes in variant classification, previously analyzed cancer-predisposition targeted sequencing data were assessed (4,929 individuals from two genetic centers) (Variant-centered reanalysis, Figure 3). This method highlighted 45 variants with major changes between the time of analysis and December 2019, that were proposed for manual review by their referring geneticists (Tables S5 and S6).

**Figure 3.**
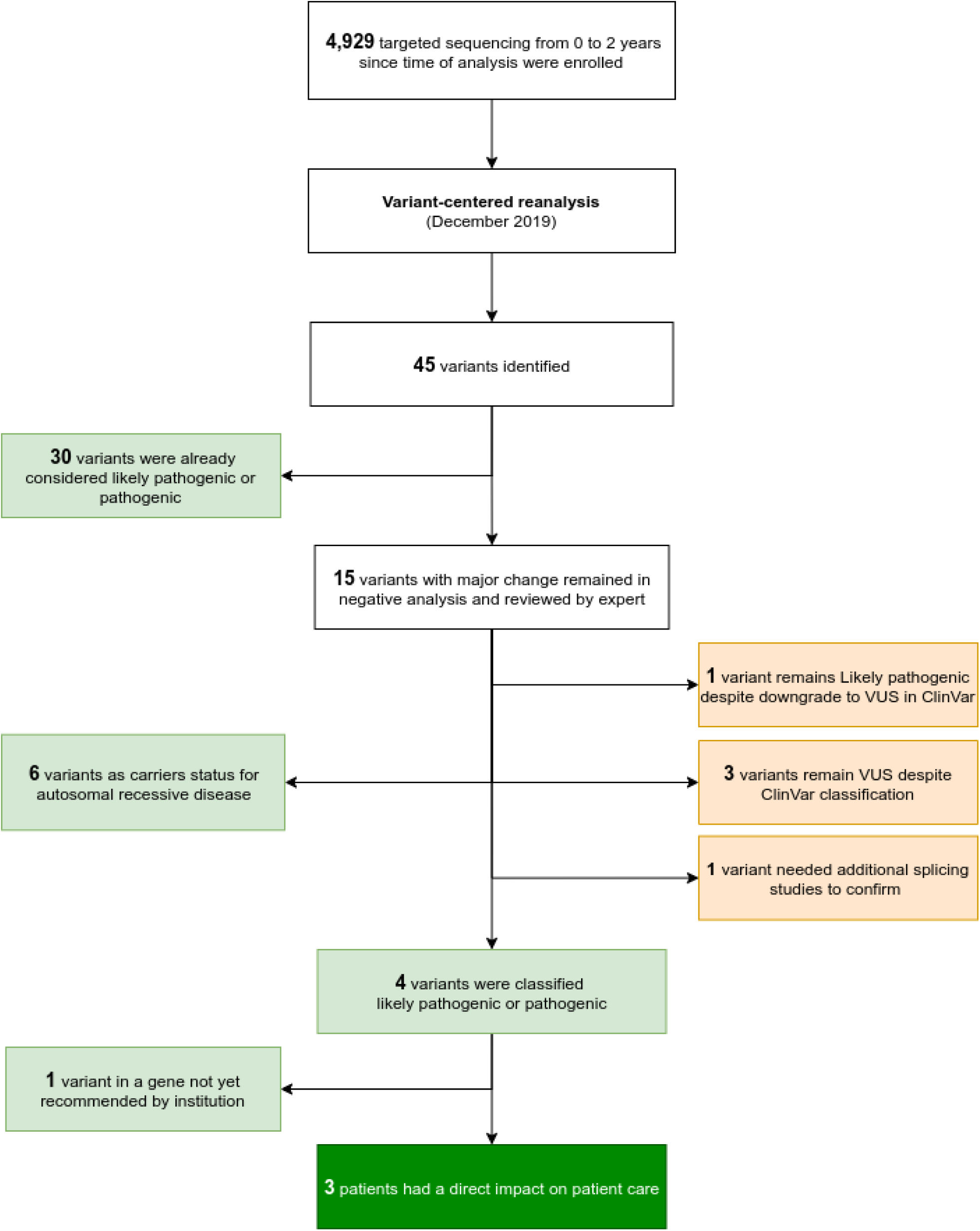
Experimental design of the variant-centered reanalysis. Flow charts describing how the sequencing data were reinterpreted according to variant reclassification only. Green: new diagnosis. Light green: confirmed variant classification. Orange: excluded variants.

Among the 45 variants, 30 had been already manually reported by the clinical geneticists as likely pathogenic or pathogenic. The 15 unreported variants were manually curated. Among them, 14 variants were newly classified as likely pathogenic or pathogenic and 1 was downgraded as a VUS in ClinVar. The manual curation of these 14 variants lead to the conclusion that: 6/14 corresponded to a carrier status for a recessive disorder, 3/14 were manually classified as VUS and 5/14 were submitted to a multidisciplinary meeting for external review. Finally, four of these latter five were classified as likely pathogenic or pathogenic by experts leading to additional diagnoses. One remained classified as a VUS and complementary studies on the patient’s mRNA were proposed before conclusion (*PALB2*, NM_024675.3:c.3350+4A>G). At last, a 89% validation rate (40 of 45) of major changes were observed. This variant reclassification tracking system allowed an additional diagnosis per 1,000 analyses.

Replication of the variant-centered reanalysis was performed in the exome sequencing cohort, looking for an exact match in variants. Selective reanalysis in previous exome sequencing analysis (1,000 individuals in 3 genomic centers) highlighted less than one variant per exome (only 297 variants) with major changes between the time of analysis and December 2019. These 297 variants were then explored by clinical geneticists. Among all 297 variants, one variant (*POLG*, NM_002693.2:c.2243G>C (p.Trp748Ser)) was automatically reclassified as pathogenic by our IQR outlier submission method, was initially reported as VUS and helped us to confirm the diagnosis. A compound heterozygosity was observed with a pathogenic variant (*POLG*, NM_002693.3:c.1399G>A (p.Ala467Thr)). Exome sequencing reanalysis with the variant-centered reanalysis provides also an additional diagnosis per 1,000 analysis.

### Monitoring ClinVar gene-disease association knowledge

A focus has been made on rarely explored gene-disease association in ClinVar data. To discriminate candidate genes from definitive gene-disease association in ClinVarome, unsupervised clustering was performed based on the following criteria (i) count of likely pathogenic and pathogenic variants, (ii) highest variant classification (iii) highest ClinVar review variant confidence and (iv) time interval between the first to the last pathogenic variant submission. According to distances between clusters and model dendrogram, the number of clusters was set to four (Figure 4). Careful observation of these clusters identified objective patterns to understand the classification. We observed that genes in the 1st and 2nd clusters had a reproducibility event in pathogenicity status, thus giving them strong confidence. Genes from the 1st-cluster hold pathogenic variants with ClinVar’s two stars or more of review confidence and the 2nd cluster genes include pathogenic variants with different entries dates and less than two stars of review confidence. Genes in the 3rd cluster got one strong argument for pathogenicity but needed another event to be fully confirmed (the 3rd cluster genes contained at least one pathogenic variant and all pathogenic entries were added at the same date). Since genes in the 4th cluster got only likely pathogenic variants, their gene-disease association remained to be confirmed (Table S7).

**Figure 4.**
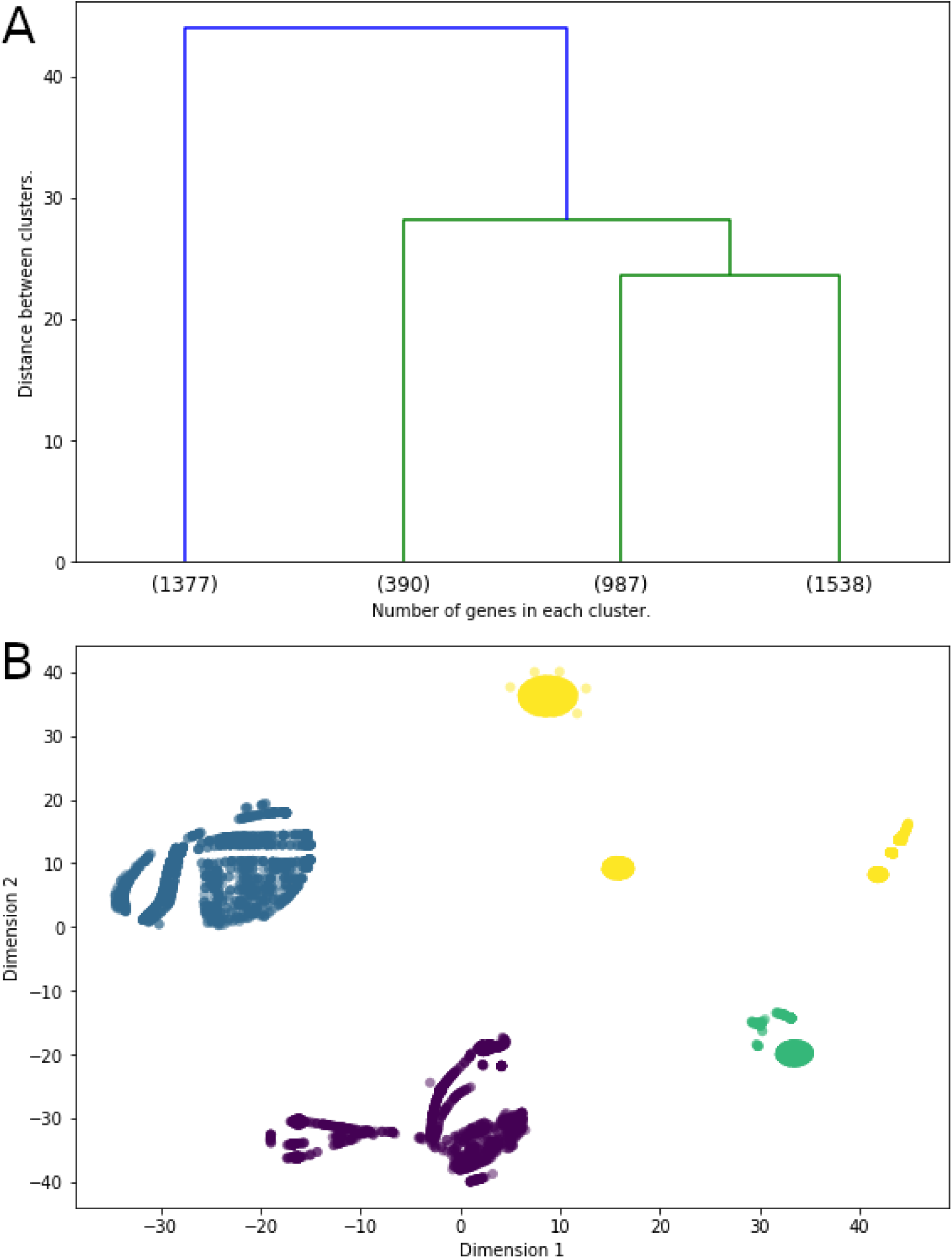
ClinVarome morbid genes exploration and gene-disease validity classification. A. Agglomerative clustering dendrogram of ClinVarome in December 2019. B. t-SNE representation of ClinVarome four variables by gene data. Green : 0 star (390 genes), Yellow : 1 star (987 genes), Blue : 2 stars (1538 genes), Purple : 3 stars (1377 genes).

To assess the exhaustivity of the ClinVarome, a comparison with the OMIM database was performed. In December 2019, there was a 95% overlap (3,675/3,858) between OMIM morbid clinical genes and ClinVarome. Overall, 365 genes were referenced only in OMIM and not in ClinVar, and 519 genes were referenced only in ClinVar and not in OMIM. The evaluation focused on these 519 specific genes to assess their potential value in additional diagnoses.

Among the 519 *ClinVarome* only genes in December 2019, 15 genes were in the 1st cluster 1 and 60 genes were in the 2nd cluster (i.e. 75 high-confidence genes), 140 genes in the 3rd cluster and 304 genes in the 4th cluster. Then, we monitored their inclusion in the OMIM morbid list in the upcoming months. Among the 519 genes, 55 were reported OMIM morbid eight months later in August 2020, including 15 of the 75 (20%) initial high-confidence genes. In the same period, 125 of the 140 OMIM morbid genes additional entries were referenced in ClinVar. This observation suggested that candidate genes in ClinVar may be considered as diagnostic genes before the OMIM validation of the gene-disease causality.

### Clinical impact of ClinVarome morbid genes not available in OMIM

We evaluated the relevance of this approach by performing a selective reanalysis of a subsample of the new entries in the ClinVarome, focusing only on the 75 genes that were absent from OMIM Morbid and referenced in ClinVarome’s first and second clusters (Gene-centered reanalysis). This experiment highlighted 42 variants in 356 negative exome sequencing data. In this dataset, 42 variants were prioritized and were proposed for further interpretation. Among them, 39 were excluded by the expert. The experts’ arguments included the presence of variants unrelated to the disease phenotype, or a single case series available in the literature. Three variants were further explored with Sanger sequencing validation, of which 2 were excluded due to artifact status or discordant inheritance pattern (Figure 5).

**Figure 5.**
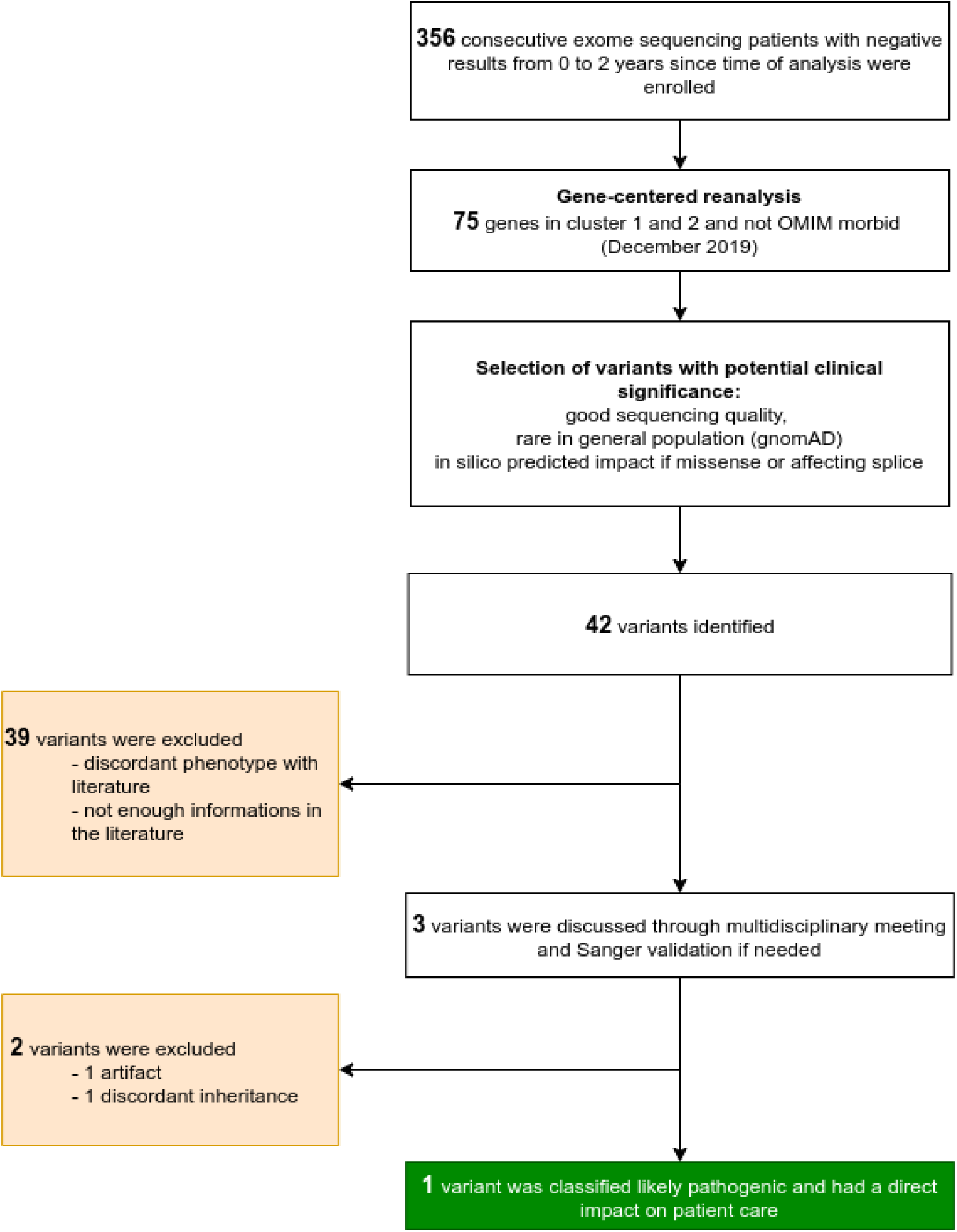
Experimental design for a targeted gene-centered reanalysis. These 75 genes were reported in ClinVarome and not OMIM, and classified as related to a disease (cluster 1 and 2). This list of 75 genes was used for the reinterpretation of negative exome sequencing data (n=346). Green: new diagnosis. Orange: excluded variants.

Overall, this method could ascertain a new diagnosis from the 356 negative exome sequencing data. A nonsense *DLG4* variant NM_001128827.1:c.1840C>T(p.Arg614Ter) was reported as likely pathogenic, responsible for the patient’s phenotype (intellectual disability and microcephaly).

## Discussion

With the increasing amount of genetic testing performed in healthcare, there is a critical need for standardized methods to enable a prospective genomic data reinterpretation in clinical routine. Through the reassessment of variant pathogenicity and gene-phenotype associations in ClinVar, Genome Alert!’s data mining method proposes the automatic report of a handful of variants that can reasonably be manually interpreted. Our method was applied to a multicentric series of 4,929 sequencing tests with various local bioinformatic systems. Genome Alert! successfully allowed new diagnoses in targeted and exome sequencing through query of laboratory’s VCFs or variant database, and proposes the first portable and open-source framework for an automated reanalysis of sequencing data.

Retrospective monitoring of the cutting-edge medical literature on existing genomic data is a major concern for paving the way to genomic medicine ^32^. There are numerous technical and medical challenges in setting up a routine procedure for reanalysis. This work explored the dynamic of change across all fields of genomic medicine in ClinVar.

Several medical indications for genomic testing were noticed to bear numerous changes in variant classification. Retrospective analysis of the ClinVar database provided an estimation of new clinically relevant information reported each month, that may lead to additional diagnoses in the existing data ^33^. Overall, 9.94 % (1,125) likely pathogenic variants were *eventually* downgraded and reclassified as benign, likely benign, unknown significance or with conflicting interpretation status in ClinVar over the study period (Table S3). This metric was contrasting with 2.98% (834) of the pathogenic variants that were reclassified. This observation is higher than a previous estimation performed in 2019, observing changes in respectively 0.46% (171 downgraded likely pathogenic variants with potential clinical impact) and 0.07% (46 downgraded pathogenic variants with potential clinical impact) ^34^. Main difference between these two studies is the selection of variants monitored. Unlike previous studies which monitor the evolution of all entries between 1 January 2016 and 1 July 2019, we focus our analysis on the evolution of variant classification available in July 2017 only. The results of the present work is close to the estimated internal consistency of current ACMG-AMP Guidelines for variant classification for likely pathogenic and pathogenic variants, respectively ranging from to 90-99% and 97-99.9% ^7^. This observation highlights the required carefulness in returning results to the families for likely pathogenic variants since such information could be used for genetic counseling and patient management.

Genome Alert! methods are based on the processing of submissions from the ClinVar full XML release. As HGVS genomic nomenclature is adopted by the community, Genome Alert! attributes a unique variant ID based on HGVSg. As such, these variants with potential clinical significance are reported by Genome Alert! should be queryable *a priori* in each genomic center. We named this method ClinVCF, a prerequisite for the variant monitoring tool, Variant Alert!.

Clinical impact of changes in variant classification (variant-centered reanalysis) provided in our targeted and exome sequencing cohort provided an additional diagnosis per 1,000 analyses. As time from initial analysis varies from 0 to 2 years, this diagnostic yield will certainly increase with time. This automated system is better for large cohorts of targeted sequencing, with a low number of variants to reinterpret and reaching 10% diagnostic yield in the re-examined variants. Recent literature emphasizes the importance of a standardized procedure adapted for sequencing data reanalysis for considering few candidate variants after an accurate annotation of new gene-phenotype associations and filtering procedure ^32^.

A particular effort was made to evaluate confidence in the reported information to reach a consensus across multiple annotations. The prospective reassessment of ClinVar highlighted numerous conflicts in variant classification, and sometimes conflicts in well-studied pathogenic variants. Although our system rarely reclassifies variants with conflicting interpretations, this automatic reclassification method aims to at least remove these potential errors. The expert-review of ClinVCF automatic reclassification validates this method based on outlier submission removal by the IQR method, and succeed to reclassify abnormalities such as the well-known *HFE* pathogenic variant NM_000410.3:c.845G>A (p.Cys282Tyr). This work highlights the value of the persistence over time of a classification for a relevant genomic information. This work specifically focused on oncogenetics and cardiogenetics, fields where variant interpretations are particularly conflicting and shifting ^35 36^. Overall in the ClinVar database, 188 variants could be reclassified in the 29-month period (ranging from 2017-2019). Eight months later, in August 2020, a total of 307 variants were reclassified, highlighting the importance of a systematic and automated variant reassessment (Figure S2).

Existing literature for gene-centered reanalysis emphasized the importance of OMIM as an updated resource. We chose to focus our reanalysis on 75 high-confidence ClinVarome morbid genes (1st and 2nd clusters) not available in OMIM Morbid. Complementary to OMIM morbid genes, these high-confidence *ClinVarome* morbid genes from the 1st and 2nd clusters could provide additional diagnoses in exome or genome sequencing analysis (gene-centered reanalysis). One additional diagnosis was identified with this tight subsampling of variants among the 356 negative exomes, validating this proof of concept. Additional experiments could be performed to fully evaluate the ClinVarome, such as a reanalysis with the full list of ClinVarome morbid genes not found in OMIM, or with additional cohorts.

The ClinVarome gene-disease validity association method was based on the Clinical Genome Resource recommendations of clinical validity of gene-disease associations evaluation ^27^. Among the proposed criteria, this work identified the four most discriminative features for gene-disease clinical validity available in ClinVar data. Overall, the evaluation relies mainly on the amount of knowledge, but also on reported review confidence and more importantly on the time-scale of entries. The Genome Alert! gene-curation via machine learning methods provides an original attempt for automated evaluation of gene confidence in disease. Genome Alert! proposes a standardized clinical validity confidence score that could allow a prospective gene-phenotype association assessment. As such, this approach could be useful to update *in silico* gene panels. This procedure proposes a complementary approach to the simple aggregation of multiple expert-reviewed databases such as DDG2P, Genomic England PanelApp or ClinGen Gene-disease validity, which are partially overlapping ^37^.

In summary, Genome Alert! highlights changes with potential clinical significance. To our knowledge, this is the first large retrospective study of an automated system for sequencing data reinterpretation. This procedure enables the systematic and reproducible reinterpretation of acquired sequencing data in a clinical routine, with a limited human resource impact and a significant diagnostic yield improvement. Genome Alert! provides an open-source accessible framework to the community, thus hoping to be applicable in every genetic center.

## Data Availability

Genome Alert! results are publicly available at https://genomealert.univ-grenoble-alpes.fr/. Relevant data used to generate Genome Alert! results are available from ClinVar FTP (all monthly ClinVar full XML release data were downloaded at https://ftp.ncbi.nlm.nih.gov/pub/clinvar/xml/) and in the following resources: OMIM (https://omim.org/), Genomic England PanelApp (https://panelapp.genomicsengland.co.uk/) and RefSeq annotation (ftp://ftp.ncbi.nlm.nih.gov/refseq/H_sapiens/annotation/GRCh38_latest/refseq_identifiers/GRCh38_latest_genomic.gff.gz). All code for generating Genome Alert! procedures are available at public GitHub repositories: ClinVCF tool for ClinVar XML full release processing and extraction to VCF format (https://github.com/SeqOne/clinvcf), Variant Alert! tool to compare ClinVCF release (https://github.com/SeqOne/variant_alert), ClinVarome tool to evaluate clinical validity of ClinVar morbid genes (https://github.com/SeqOne/clinvarome) and the Genome Alert! shiny app (https://github.com/SeqOne/GenomeAlert_app).

https://ftp.ncbi.nlm.nih.gov/pub/clinvar/xml/

https://github.com/SeqOne/clinvcf

https://github.com/SeqOne/variant_alert

https://github.com/SeqOne/clinvarome

https://github.com/SeqOne/GenomeAlert_app

## Data and Code Availability

### Software summary

Project name: Genome Alert!

Project home page: https://genomealert.univ-grenoble-alpes.fr/

Operating system(s): UNIX (Mac, Linux)

Programming language: Nim, Python, R

License: Apache Licence 2.0

Any restrictions to use by non-academics: No

## Acknowledgments

We sincerely thank all patients, clinicians, biologists and bioinformaticians involved in this project. A special thanks to Virginie Bernard and Valentin Klein for their feedback and to external reviewers through the publication process. This work has been partially supported by MIAI@Grenoble Alpes (ANR-19-P3IA-0003).

## Web-resources

Genome Alert! webapp, https://genomealert.univ-grenoble-alpes.fr/

ClinVCF, https://github.com/SeqOne/clinvcf

Variant Alert!, https://github.com/SeqOne/variant_alert

ClinVarome, https://github.com/SeqOne/clinvarome

Genome Alert! Shiny app, https://github.com/SeqOne/GenomeAlert_app

ClinVar, https://www.ncbi.nlm.nih.gov/clinvar/

OMIM, https://www.omim.org/

Genomic England Panel App, https://panelapp.genomicsengland.co.uk/

## Ethics declarations

### Ethics approval and consent to participate

Patients referred to the Eurofins Biomnis laboratory, Cerba laboratory and CHU de Rouen Molecular Genetics laboratory provided written consent for analysis of their DNA by NGS including research analysis for the purpose of obtaining a molecular diagnosis. Sequencing samples were anonymized. This research conforms to the principles of the Helsinki Declaration.

### Consent for publication

Not applicable.

## Authors’ contributions

K.Y., F.L., S.Ca., D.B., A-L.B., J.A., G.N., J.T., N.P. designed the study; K.Y., F.L., S.B.D., D.T., A.B., A.L., J-M.C., V.G., L.R., P.R., N.P. collected data; K.Y., S.Co., M.B., R.L., Q.F., A.D., N.S., S.B., J.A. coded the bioinformatics tools; K.Y., F.L., S.B.D., D.T, A.B., A.L., J-M.C, V.G., L.R., P.R, S.C., Q.F., J.A. performed the analysis ; K.Y., F.L., Q.F., Q.T., J-M.H., D.B., G.N, J.T., N.P. wrote the draft. A-L.B., J.A., D.G., L.M., G.N, J.T., N.P. supervised the study. All authors reviewed and edited the drafts.

