## Supplementary Table and Figures for "Genome Alert!: a standardized procedure for genomic variant reinterpretation and automated genotype-phenotype reassessment in clinical routine"

**Supplemental Tables**

Table S1. Genome Alert! Clinical impact change status definition.

Table S2. Monitoring of variant classification specific for Genomics England PanelApp panels from July 2017 to December 2019.

Table S3. Variant classification evolution in December 2019 among 144.943 variants available in June 2017.

Table S4. List of reviewed variants by clinical experts for validation of Genome Alert! automatic reclassification of conflicting interpretation of pathogenicity status.

Table S5. Genome Alert! variant classification tracking system analysis of laboratory 1 variant database from the oncogenetic targeted sequencing panel.

Table S6. Genome Alert! variant classification tracking system analysis of oncogenetic panels from laboratory 2.

Table S7. Median per cluster of the four variables used to classify gene-disease clinical validity.

Table S1. Genome Alert! Clinical impact change status definition.

| **Previous** | **New** | **Clinical impact** |
| --- | --- | --- |
| **Variant** |  |  |
| Absent | .. | null |
|  | Benign and/or likely_benign | null |
|  | Uncertain_significance or Conflicting_interpretations_of_pathogenicity | unknown |
|  | Likely_pathogenic | major: new pathogenicity |
|  | Pathogenic | major: new pathogenicity |
| Benign and/or likely_benign | .. | deleted from database |
|  | Uncertain_significance or Conflicting_interpretations_of_pathogenicity | unknown |
|  | Likely_pathogenic | major: new pathogenicity |
|  | Pathogenic | major: new pathogenicity |
| Uncertain_significance or  Conflicting_interpretations_of_pathogenicity | .. | deleted from database |
|  | Benign and/or likely_benign | minor |
|  | Likely_pathogenic | major : new pathogenicity |
|  | Pathogenic | major : new pathogenicity |
| Likely_pathogenic or Pathogenic/Likely_pathogenic | .. | deleted from database |
|  | Benign and/or likely_benign | major: revoked pathogenicity |
|  | Uncertain_significance or Conflicting_interpretations_of_pathogenicity | major : revoked pathogenicity |
|  | Pathogenic | minor |
| Pathogenic | .. | deleted from database |
|  | Benign and/or likely_benign | major: revoked pathogenicity |
|  | Uncertain_significance or Conflicting_interpretations_of_pathogenicity | major: revoked pathogenicity |
|  | Likely_pathogenic | minor |
| **Gene** |  |  |
| .. | NEW_PATHOGENICITY | major : new pathogenicity |
| Any | LOST_PATHOGENICITY | major: revoked pathogenicity |

Table S2. Monitoring of variant classification specific for Genomics England PanelApp panels from July 2017 to December 2019.
In Supplementary Excel spreadsheets.

Table S3. Variant classification evolution in December 2019 among 144.943 variants available in July 2017. Perc. change category = percentage of variant classification changes by ACMG class.

| **Old classification** | **type** | **count** | **Initial number** | **Number of changes** | **Percentage changes** | **Perc. change category** |
| --- | --- | --- | --- | --- | --- | --- |
| *Benign* | warning | 59 | 14255 | 1502 | 10.54 | 0.41 |
|  | Likely benign | 1262 |  |  |  | 8.85 |
|  | Uncertain significance | 180 |  |  |  | 1.26 |
|  | Likely pathogenic | 0 |  |  |  | 0.00 |
|  | Pathogenic | 1 |  |  |  | 0.01 |
| *Likely benign and Benign/Likely benign* | warning | 38 | 31612 | 2658 | 8.41 | 0.12 |
|  | Benign | 578 |  |  |  | 1.83 |
|  | Uncertain significance | 2038 |  |  |  | 6.45 |
|  | Likely pathogenic | 2 |  |  |  | 0.01 |
|  | Pathogenic | 2 |  |  |  | 0.01 |
| *Uncertain significance and Conflicting interpretations of pathogenicity* | warning | 555 | 59683 | 2157 | 3.61 | 0.93 |
|  | Benign | 185 |  |  |  | 0.31 |
|  | Likely benign | 962 |  |  |  | 1.61 |
|  | Likely pathogenic | 350 |  |  |  | 0.59 |
|  | Pathogenic | 105 |  |  |  | 0.18 |
| *Likely pathogenic and Pathogenic/Likely pathogenic* | warning | 23 | 11417 | 1513 | 13.25 | 0.20 |
|  | Benign | 1 |  |  |  | 0.01 |
|  | Likely benign | 2 |  |  |  | 0.02 |
|  | Uncertain significance | 1109 |  |  |  | **9.71** |
|  | Pathogenic | 378 |  |  |  | 3.31 |
| *Pathogenic* | warning | 246 | 27976 | 2429 | 8.68 | 0.88 |
|  | Benign | 9 |  |  |  | 0.03 |
|  | Likely benign | 5 |  |  |  | 0.02 |
|  | Uncertain significance | 574 |  |  |  | 2.05 |
|  | Likely pathogenic | 1595 |  |  |  | 5.70 |

Table S4. List of reviewed variants by clinical experts for validation of Genome Alert! automatic reclassification of conflicting interpretation of pathogenicity status.

| **Conflicting variants reclassified as (likely) pathogenic - December 2019** | **Reclassification confidence** | **Expert classification** |
| --- | --- | --- |
| NM_170707.4(LMNA):c.725C>T (p.Ala242Val) | 1 | Class 5 |
| NM_000258.2(MYL3):c.427G>A (p.Glu143Lys) | 1 | Class 4 |
| NM_007078.3:(LDB3)c.690-4678C>T,NM_001080116.1(LDB3):c.494C>T (p.Ala165Val) | 1 | Class 4 |
| NM_000256.3(MYBPC3):c.3815-1G>A | 1 | Class 5 |
| NM_000256.3(MYBPC3):c.2905+1G>A | 1 | Class 5 |
| NM_000256.3(MYBPC3):c.1504C>T (p.Arg502Trp) | 3 | Class 3 or 4 |
| NM_000257.4(MYH7):c.2011C>T (p.Arg671Cys) | 1 | Class 5 |
| NM_000257.4(MYH7):c.1324C>T (p.Arg442Cys) | 1 | Class 5 |
| NM_000257.4(MYH7):c.728G>A (p.Arg243His) | 1 | Class 5 |
| NM_001018005.2(TPM1):c.644C>T (p.Ser215Leu) | 1 | Class 4 |
| NM_024422.6(DSC2):c.2125+1del | 1 | Class 5 |
| NM_001048174.2(MUTYH):c.849+3A>C | 3 | Class 5 |
| NM_001128425.1(MUTYH):c.820C>T (p.Arg274Trp) | 1 | Class 3 |
| NM_001048171.1(MUTYH):c.267G>A (p.Trp89Ter) | 3 | Class 4 |
| NM_000179.2(MSH6):c.1109T>C (p.Leu370Ser) | 1 | Class 3 |
| NM_000179.2(MSH6):c.1295T>C (p.Phe432Ser) | 2 | Class 3 |
| NM_000179.2(MSH6):c.3725G>A (p.Arg1242His) | 2 | Class 4 |
| NM_000535.7(PMS2):c.2521del (p.Trp841fs) | 1 | Class 4 |
| NM_001126112.2(TP53):c.646G>A (p.Val216Met) | 1 | Class 5 |
| NM_001126112.2(TP53):c.374C>T (p.Thr125Met) | 2 | Class 5 |

Table S5. Genome Alert! variant classification tracking system analysis of laboratory 1 variant database from the oncogenetic targeted sequencing panel.
In Supplementary spreadsheets.

Table S6. Genome Alert! variant classification tracking system analysis of oncogenetic panels from laboratory 2.
In Supplementary spreadsheets.

Table S7. Median per cluster of the four variables used to classify gene-disease clinical validity.

| **cluster name** | **Highest ACMG classification found in a variant per gene** | **Highest ClinVar review confidence (from 0 to 4 stars) found in likely pathogenic or pathogenic variants per gene** | **Time interval between the first entry and last entry of a likely pathogenic or pathogenic variant per gene** | **Count of likely pathogenic or pathogenic variants per gene** |
| --- | --- | --- | --- | --- |
| 4th | **4** | 1 | 0 | 1 |
| 3rd | **5** | 0 | **0** | 2 |
| 2nd | 5 | 1 | **19** | 7 |
| 1st | 5 | **2** | 26 | 24 |

**Supplemental Figures**
Figure S1. Automatic reclassification of variants with conflicting interpretations of pathogenicity workflow from July 2017 to December 2019 and August 2020
Figure S2. Evolution of conflicting interpretation of pathogenicity status variants reclassified by Genome Alert! from December 2017 and December 2020.

Figure S1. Automatic reclassification of variants with conflicting interpretations of pathogenicity workflow from July 2017 to December 2019 and August 2020

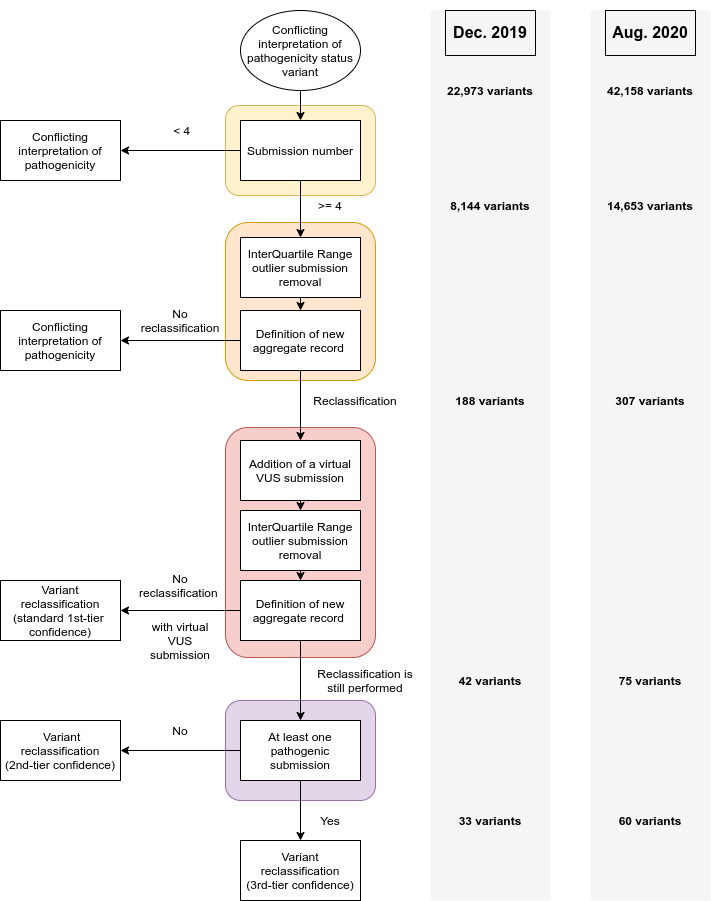

Figure S2. Evolution of conflicting interpretation of pathogenicity status variants reclassified by Genome Alert!.

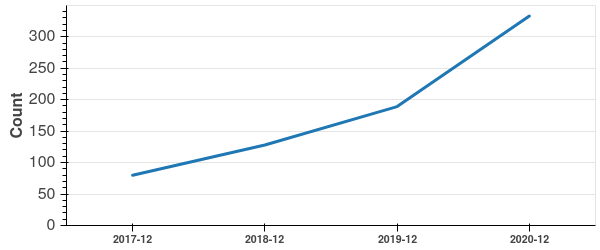
